# Comparative Evaluation of Microstructural Diffusion Methods in Characterizing Multiple Sclerosis Lesions: The Importance of multi-b shells acquisition

**DOI:** 10.64898/2026.03.15.26348428

**Authors:** Chaoyang Jin, Ahmad Tubasi, Ke Xu, Caroline Gheen, Taegan Vinersky, Hakmook Kang, Xiaoyu Jiang, Francesca Bagnato, Junzhong Xu

## Abstract

**Background:** Diffusion MRI (dMRI) is widely used to assess microstructural abnormalities in multiple sclerosis (MS), yet conventional diffusion tensor imaging (DTI) is limited by single b-shell acquisitions and reduced pathological specificity. Higher-order diffusion models enabled by multi-b-shell data may provide complementary information, but their relative performance across tissue classes remains unclear.

**Purpose:** To evaluate lesion-resolved microstructural alterations across MS tissue classes using multiple diffusion models and to assess the impact of diffusion acquisition strategy on discriminative performance.

**Methods:** Multi-shell dMRI was acquired in 57 treatment-naïve patients with early MS and 17 healthy controls. Five diffusion models were evaluated (DTI, DKI, NODDI, SMT, and SMI). 3602 manually delineated ROIs, including chronic black holes, T2 lesions, lesion-matched normal-appearing white matter (NAWM), and normal white matter (NWM), were analyzed. Microstructural differences were assessed using linear mixed-effects models, and discriminative performance was evaluated using ROC analysis across single-shell, multi-shell, and joint modeling strategies. Feature selection was performed using LASSO regression.

**Results:** Across all models, lesions exhibited coherent microstructural abnormalities relative to normal white matter, while NAWM showed concordant but more subtle alterations. Lesion–normal tissue contrasts demonstrated strong discriminative performance, whereas classification of NAWM versus NWM and lesion subtypes remained limited, reflecting substantial biological overlap. Two b-shell and joint modeling approaches consistently outperformed single-shell analyses, yielding the highest AUCs. LASSO identified a small set of biologically meaningful diffusion features driving tissue discrimination.

**Conclusion:** Multi-b-shell diffusion MRI enables more robust and informative characterization of MS-related white matter pathology than single-shell acquisitions alone, supporting multi-model, multi-b-shell strategies for lesion-resolved assessment in MS.

## Introduction

Multiple sclerosis (MS) is an immune-mediated disease of the central nervous system characterized by inflammatory activity, demyelination, and progressive axonal damage[1]. Although the clinical course of MS is often relapsing in nature, neuroaxonal degeneration is now understood to occur early and to continue throughout disease progression[2, 3]. Magnetic resonance imaging (MRI) plays a central role in the diagnosis and longitudinal monitoring of MS, with conventional T1- and T2-weighted sequences commonly used to detect focal lesions and global tissue loss[4, 5]. Acute inflammatory activity is visualized as gadolinium-enhancing lesions, which typically evolve into chronic T2-hyperintense abnormalities; a subset of these lesions further develops into persistent T1-hypointense chronic black holes (cBHs), reflecting severe axonal injury [6]. Despite their widespread clinical use, conventional MRI markers lack pathological specificity and exhibit limited sensitivity to microstructural changes, contributing to the well-known dissociation between imaging findings and clinical disability[7]. This limitation largely arises from the inability of standard MRI techniques to capture subtle and diffuse tissue damage within normal-appearing white matter and gray matter, underscoring the need for advanced imaging biomarkers capable of quantitatively characterizing neurodegenerative processes in MS[8, 9].

Diffusion MRI (dMRI) is widely used to probe microstructural abnormalities in multiple sclerosis (MS), including diffuse injury in normal-appearing white matter that may be inconspicuous on conventional MRI[10, 11]. Conventional diffusion tensor imaging (DTI) provides sensitive markers of tissue alteration and has been extensively applied in MS research and clinical studies[12]. However, tensor-derived metrics are inherently limited in pathological specificity because they represent compartment-averaged signals and can be confounded by complex fiber geometry, partial volume effects, and microstructural heterogeneity within lesions[13]. As a result, DTI metrics may reflect a mixture of underlying pathological processes rather than specific microstructural alterations[14, 15].

Many limitations of DTI arise not only from the tensor model itself but also from the underlying diffusion acquisition strategy[16]. Most clinical diffusion MRI protocols rely on single b-shell acquisitions, which restrict analyses to tensor-based models and reduce sensitivity to complex tissue microstructure[17, 18]. In contrast, multi-b-shell acquisitions enable the application of higher-order diffusion models that provide more biologically interpretable microstructural information[19]. Using such data, approaches including diffusion kurtosis imaging (DKI), neurite orientation dispersion and density imaging(NODDI), the spherical mean technique (SMT), and standard model imaging (SMI) have been applied in MS to characterize tissue alterations beyond conventional DTI[20–23]. However, their relative sensitivity, interpretability, and robustness across different tissue compartments and lesion phenotypes remain incompletely understood. In addition, most prior studies have relied on subject-level averaging or automated atlas-based analyses, limiting their ability to capture the pronounced microstructural heterogeneity within MS lesions[24–27].

In this study, we performed a systematic evaluation of five diffusion modeling frameworks, including conventional DTI and the higher-order models DKI, NODDI, SMT, and SMI, with a particular focus on the role of diffusion acquisition strategy. To enable lesion-resolved analyses, we examined more than 3,000 manually delineated regions of interest from treatment-naïve individuals at early stages of MS, spanning five white matter tissue categories that included normal-appearing tissue and distinct lesion subtypes. Diffusion-derived metrics were compared across tissue classes at both the individual parameter and model levels, with explicit comparisons of single b-shell, multi-b-shell, and joint modeling strategies. We hypothesized that multi-b-shell acquisitions would enable diffusion metrics to capture graded microstructural differences across MS tissue classes and that higher-order diffusion models would provide complementary and more discriminative information relative to conventional DTI for characterizing MS-related white matter pathology.

## Methods

### Study design and cohort

The study protocol was approved by the Institutional Review Boards of Vanderbilt University Medical Center and the Nashville VA Medical Center, and all procedures were conducted in accordance with the Declaration of Helsinki. Written informed consent was obtained from all participants.

This study included 57 patients with newly diagnosed multiple sclerosis and 17 age-matched healthy controls. MS participants underwent clinical assessment using the Expanded Disability Status Scale (EDSS), the Timed 25-Foot Walk (T25FW), and the 9-Hole Peg Test (9HPT)[28–30]. Cognitive function was evaluated using the Minimal Assessment of Cognitive Function in Multiple Sclerosis (MACFIMS) battery, including the Symbol Digit Modalities Test (SDMT), the California Verbal Learning Test (CVLT), the Brief Visuospatial Memory Test Revised (BVMT-R), the Paced Auditory Serial Addition Test (PASAT), and the Benton Judgment of Line Orientation (BJLO)[31, 32]. Exclusion criteria included contraindications to MRI, prior ischemic or hemorrhagic stroke, other systemic or central nervous system autoimmune disorders, active neoplastic or infectious disease, uncontrolled hypertension or diabetes mellitus, clinically significant cardiac disease, prior exposure to MS disease-modifying therapies except for glucocorticoids used for acute relapse management, and clinical changes occurring between neurological assessment and MRI acquisition.

### MRI acquisition

Brain MRI was acquired on a 3.0 Tesla Ingenia CX system (Philips Healthcare, Best, The Netherlands) using a volume transmit coil in combination with a 32-channel receive head coil (NOVA Medical, Wilmington, MA, USA). The imaging protocol consisted of axial T1-weighted turbo spin echo (T1w TSE), axial T2-weighted fluid-attenuated inversion recovery (T2-FLAIR), and multi-shell diffusion MRI acquisitions. For patients, contrast-enhanced imaging was performed to evaluate gadolinium-enhancing lesions, using either pre- and post-contrast three-dimensional magnetization-prepared rapid acquisition gradient echo (3D MPRAGE) sequences or post-contrast T1-weighted TSE acquisitions. All imaging sequences provided whole-brain coverage. Acquisition parameters for all sequences are summarized in Table 1.

**Table 1.** Pulse Sequence Parameters:

|  | T<sub>1</sub>-wTSE | T<sub>2</sub>-w FLAIR | DTI |
| --- | --- | --- | --- |
| Acquisition time (minutes) | ~3 | ~5 | ~8 |
| Compressing SENSE | 1.80 | None | 2 |
| Field of view(cm) | 21 × 17 × 12.8 | 21 × 17 × 12.8 | 21 × 20.2 × 12.8 |
| Acquired voxel size(mm) | 1 × 1 × 2 | 1 × 1 × 2 | 2(istropic) |
| Echo time (msec) | 125 | 9.4 | 90 |
| Inversion time (seconds) | NA | NA | NA |
| Repetition time (seconds) | 11 | 0.6-0.7 | 4.5 |
| Number of b-shells | NA | NA | 2(b = 711 seconds/mm<sup>2</sup> with 32 directions and b = 2855 seconds/mm<sup>2</sup> with 64 directions) |
| Multi-band factor | NA | NA | 2 |

NA = not applicable; T1-w TSE = T1-weighted turbo spin echo; T2-w FLAIR = T2-weighted fluid attenuated inversion recovery image; DTI = diffusion tensor imaging;

### Image analysis

#### dMRI pre-processing

For each participant, diffusion tensor imaging data were corrected for Gibbs ringing artifacts using the mrdegibbs tool from MRtrix3[33]. Susceptibility-induced geometric distortions and eddy current–related artifacts were subsequently corrected using the topup and eddy tools implemented in the FMRIB Software Library (FSL)[34].

#### Diffusion modeling and parameter estimation

Diffusion models evaluated in this study are summarised in Fig. 1. DTI metrics were derived from single-shell diffusion MRI data using MRtrix3 (https://www.mrtrix.org/). In contrast, DKI, SMT, NODDI and SMI were estimated from multi-shell diffusion MRI data using a MATLAB-based DKI estimator(https://www.mathworks.com/matlabcentral/fileexchange/65487-diffusion-kurtosis-imaging-estimator), an open-source SMT toolbox (https://github.com/ekaden/smt), the NODDI MATLAB toolbox (http://mig.cs.ucl.ac.uk/index.php?n=Tutorial.NODDImatlab) and the NYU Diffusion MRI Group SMI toolbox (https://github.com/NYU-DiffusionMRI/SMI), respectively. For all modeling approaches, voxel-wise parametric maps were generated for subsequent analyses. No spatial smoothing or denoising was applied after model fitting to preserve the native spatial resolution of the diffusion-derived maps.

**Figure 1.**
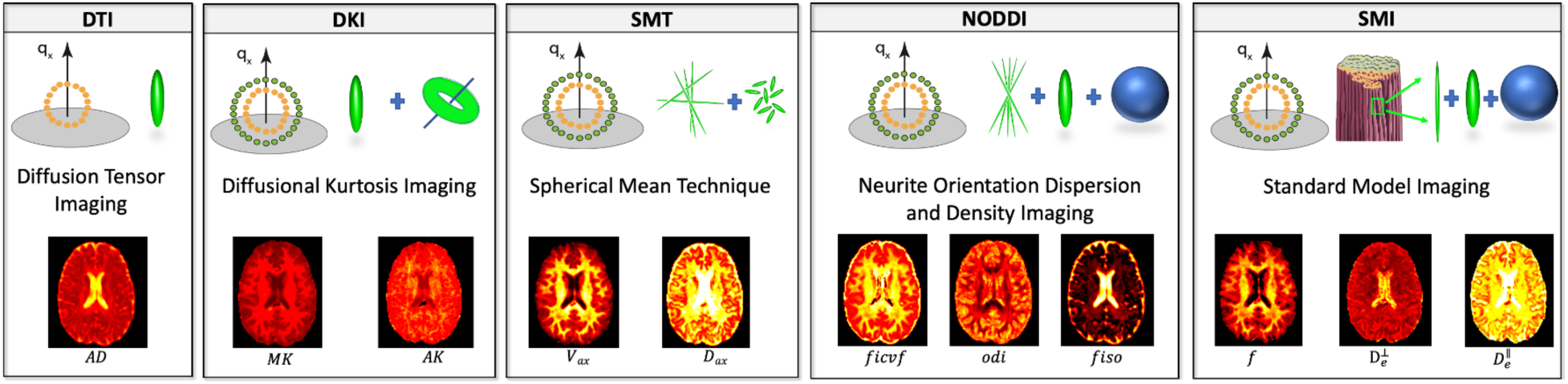
Overview of diffusion MRI models evaluated in this study. Schematic illustration of the diffusion modeling frameworks, including DTI, DKI, SMT, NODDI and SMI. The diagram highlights key modeling assumptions and acquisition requirements, with DTI based on single-shell diffusion MRI and the remaining models estimated from multi-shell data. Representative voxel-wise parametric maps are shown in the lower panel to illustrate model-dependent contrast across white matter. *AD*, axial diffusivity; *MK*, mean kurtosis; *AK*, axial kurtosis; *V*_*ax*_intra-neurite volume fraction; *D*_*ax*_, intra-neurite axial diffusivity; *ficvf*, intracellular volume fraction; *odi*, orientation dispersion index; fiso, isotropic volume fraction; *f*, intra-axonal volume fraction; 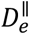 and 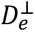, extra-axonal perpendicular and parallel diffusivities.

#### Image registration

All structural images and diffusion-derived parametric maps, including T1-weighted TSE images and maps derived from DTI, DKI, SMT, NODDI, and SMI models, were spatially registered to the corresponding T2-weighted FLAIR image using an affine transformation implemented with the FSL Linear Image Registration Tool (FLIRT). This registration approach ensured consistent anatomical alignment across imaging modalities and has been validated in our previous studies[26, 35].

#### Image analysis

Lesions and non-lesional regions of interest (ROIs) were manually delineated using Medical Imaging Processing, Analysis, and Visualization (MIPAV; version 7.3; https://mipav.cit.nih.gov/). Gadolinium-enhancing lesions were first identified on post-contrast T1-weighted images by a senior neuroradiologist (F.B., >20 years of experience) and excluded from further analysis. All T2 lesions and chronic black holes (cBHs) were subsequently segmented by a postdoctoral research fellow with a medical degree (A.T.). A matched-ROI strategy was applied to account for anatomical variability. In patients, normal-appearing white matter (NAWM) ROIs were defined relative to each lesion, including proximal NAWM located immediately anterior and posterior to the lesion and distant NAWM located in the contralateral hemisphere. In healthy controls, anatomically corresponding normal white matter ROIs were drawn to match lesion-related or NAWM locations in patients. All ROI masks were reviewed by the senior neuroradiologist (F.B.) for accuracy. Validated binary masks were then applied to co-registered parametric maps derived from DTI, DKI, SMT, NODDI, and SMI to extract regional mean values (Fig. 2).

**Figure 2.**
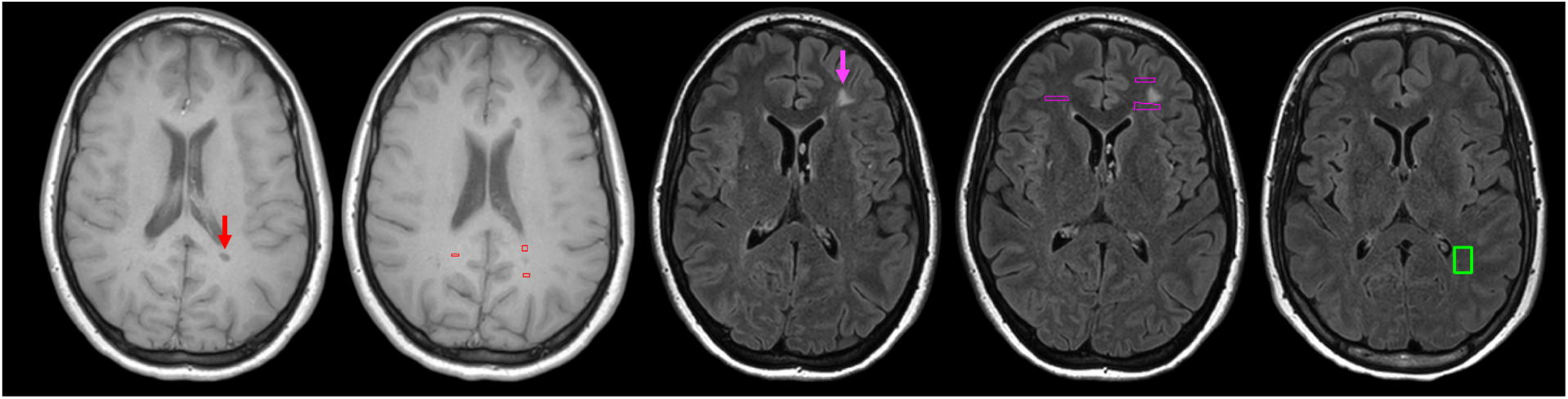
Representative examples of lesion identification and region of interest (ROI) placement. The figure displays the segmentation strategy across T1-weighted and T2-FLAIR sequences: (A) T1-weighted image showing a chronic black hole(cBHs), indicated by a red arrow. (B) T1-weighted image illustrating cBH-associated NAWM ROIs (red boxes), placed proximally to the lesions. (C) T2-FLAIR image displaying a T2-hyperintense lesion, indicated by a pink arrow. (D) T2-FLAIR image showing lesion-associated NAWM ROIs (purple boxes), placed proximally to the T2-hyperintensities. (E) T2-FLAIR image showing a normal white matter (NWM) ROI (green box) in a healthy control. Abbreviations: cBHs, chronic black holes; NAWM, normal-appearing white matter; NWM, normal white matter.

#### Statistical Analyses

Voxel-wise mean values of diffusion parameters derived from five diffusion models (DTI, DKI, SMT, NODDI, and SMI) were extracted from predefined white matter regions of interest, including cBHs, T2-lesions, cBHs-NAWM, T2-NAWM, and T2-NWM. Statistical analyses were conducted at the ROI level using mixed-effects modeling to account for within-subject dependence. Linear mixed-effects models were used to assess microstructural differences across tissue classes, with tissue type specified as a fixed effect and subject as a random intercept[36]. Six predefined pairwise contrasts were evaluated (cBHs vs. T2-NWM, T2-lesions vs. T2-NWM, cBHs vs. cBHs-NAWM, T2-lesions vs. T2-NAWM, T2-NAWM vs. T2-NWM, and cBHs vs. T2-lesions), and multiple comparisons across diffusion parameters were controlled using false discovery rate (FDR) correction.

Discriminative performance was further evaluated using receiver operating characteristic (ROC) analysis within a generalized linear mixed-effects modeling framework. Classification analyses were performed at both the individual parameter and model levels, enabling comparison of diffusion models as well as acquisition strategies, including single-shell, multi-shell, and joint modeling approaches. Within the joint modeling framework, least absolute shrinkage and selection operator (LASSO) regression was applied to identify the most discriminative diffusion parameters for each tissue contrast.

## Results

### 3.1 Participants Characteristics

A total of 57 MS patients and 17 healthy controls (HCs) were included in this study. The mean age of MS patients was 38.82 ± 11.70 years, and that of HCs was 36.94 ± 12.10 years. There was no significant difference in age between the two groups (Welch’s t-test, p = 0.577). In terms of sex distribution, the MS group included 39 females and 18 males, while the HC group included 11 females and 6 males. There was no significant difference in sex distribution between groups (χ² test, p = 0.774).

### 3.2 Differences in Diffusion MRI Model Parameters Across Tissue Classes

In total, more than 3,602 manually delineated regions of interest were included in the present study, comprising 223 cBHs, 270 cBHs-NAWM, 1,621 T2-lesions, 1,196 T2-NAWM, and 292 T2-NWM.

#### 3.2.1 Lesions and NWM

For the comparison between cBHs and NWM, widespread and significant differences were observed across diffusion metrics derived from all evaluated models, including DTI, DKI, SMT, NODDI, and SMI (Fig. 3). For the comparison between cBHs and T2-NWM, consistent patterns of microstructural alteration were observed across diffusion modeling frameworks. Specifically, DTI parameters exhibited reduced FA with increased MD, AD, and RD; DKI metrics (AK, MK, RK) were decreased; SMT showed reduced *V*_*ax*_with elevated *D*_*ex*_; NODDI revealed decreased *ficvf* and *odi* along with increased *fiso* and *Kappa*; and SMI demonstrated reduced *f* and *p*_2_ with increased *D*_*a*_, 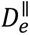 and 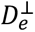. A similar pattern of alterations was also found when comparing T2-lesions to T2-NWM, indicating consistent microstructural disruption across tissue classes. Detailed statistical results, including effect sizes, confidence intervals, and corrected p-values, are provided in Supplementary Table 3.

**Figure 3.**
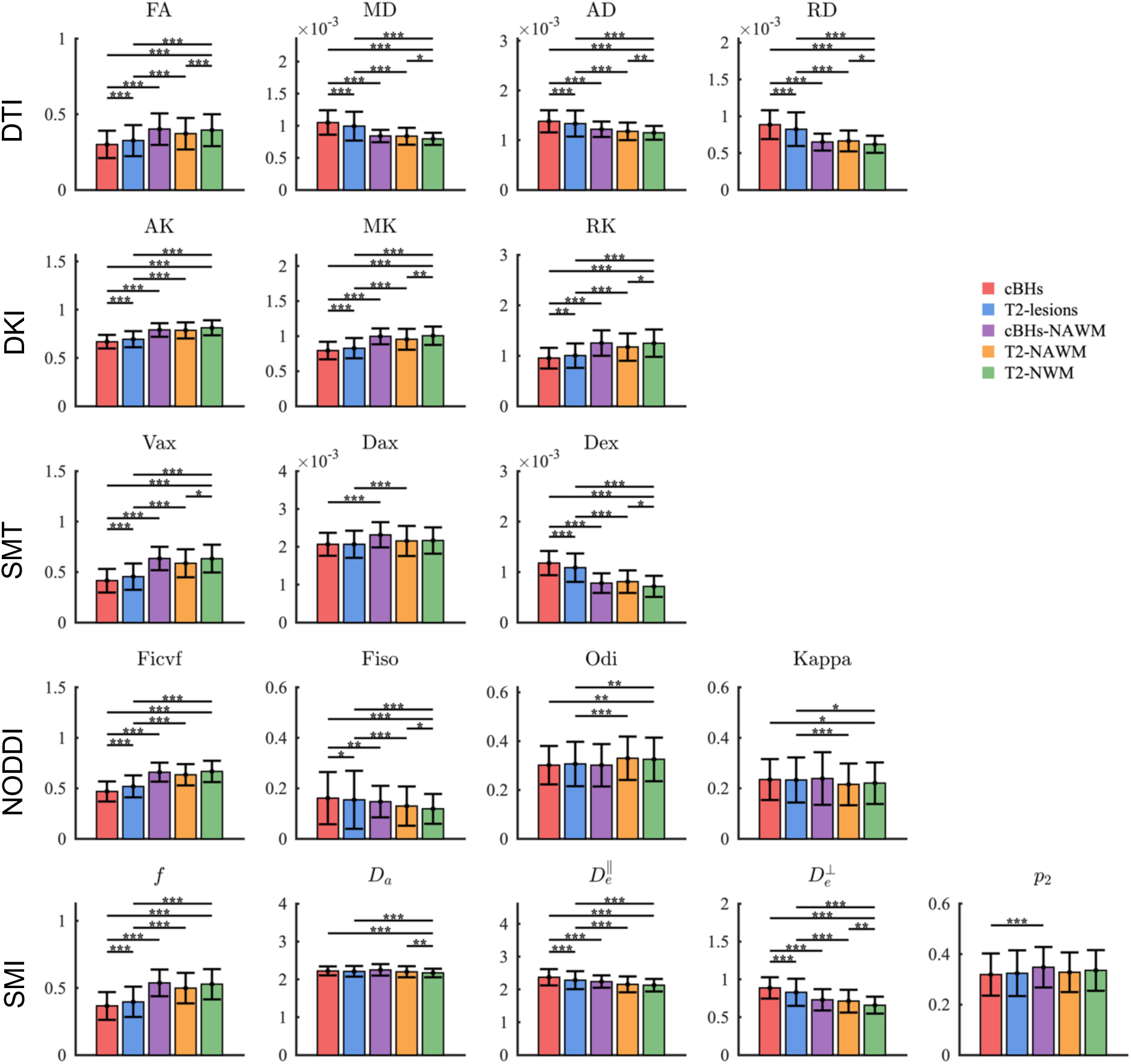
Group comparisons of diffusion metrics across five white matter tissue types in MS and HC. Tissue classes included cBHs (chronic black holes), T2-lesions, cBHs-NAWM and T2-NAWM (normal-appearing white matter), and NWM (normal white matter). Diffusion metrics were derived from DTI (fractional anisotropy [FA], mean diffusivity [MD], axial diffusivity [AD], radial diffusivity [RD]), DKI (axial kurtosis [AK], mean kurtosis [MK], radial kurtosis [RK]), SMT (intra-axonal signal fraction [*V*_*ax*_], extra-axonal diffusivity [*D*_*ex*_]), NODDI (intracellular volume fraction [*ficvf*], isotropic volume fraction [*fiso*], orientation dispersion index [*odi*], *kappa*), and SMI (intra-axonal fraction [*f*], intra-axonal diffusivity [*D*_*a*_], extra-axonal parallel diffusivity [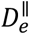], extra-axonal perpendicular diffusivity [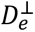], fiber orientation coherence [*p*_2_]).

#### 3.2.2 Lesions and NAWM

When comparing lesions with NAWM, the overall patterns of diffusion alterations were largely consistent with those observed in the lesions versus NWM contrast, with only a few parameters showing subtle differences. Specifically, in the SMT model, both cBHs and T2-lesions exhibited significantly reduced *D*_*ax*_relative to their respective NAWM, whereas *p*_2_ was significantly lower in cBHs but not altered in T2-lesions. For all other parameters examined, cBHs and T2-lesions showed consistent directional changes relative to NAWM (Fig. 3; Supplementary Table 3).

#### 3.2.3 NAWM and NWM

Relative to T2-NWM, T2-NAWM exhibited coherent microstructural changes consistent with subtle tissue disruption: in the DTI model, anisotropy was reduced while overall diffusivity was increased; DKI metrics (MK, RK) were decreased. SMT showed lower intra-axonal signal fraction with elevated extra-axonal diffusivity; NODDI indicated increased isotropic signal fraction; and SMI revealed elevated intra-axonal and extra-axonal diffusivities (both *D*_*a*_, and 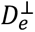).

#### 3.2.4 cBHs and T2-lesions

Across all diffusion models, cBHs differed significantly from T2-lesions. cBHs exhibited reduced anisotropy and increased diffusivity in DTI; decreased kurtosis in DKI; lower *V*_*ax*_ and higher *D*_*ex*_ in SMT; reduced *ficvf* and increased *fiso* in NODDI; and lower *f* with higher 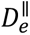 and 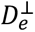 in SMI.

### 3.3 Discriminative Performance of Diffusion Model Parameters Across Tissue Classes

#### 3.3.1 Lesions and NWM

For both cBHs vs T2-NWM and T2-lesions vs T2-NWM comparisons, many diffusion parameters across all evaluated models demonstrated strong discriminative performance. In particular, high classification performance was observed for DTI metrics MD, AD, and RD; DKI metrics AK, MK, and RK; SMT measures *V*_*ax*_ and *D*_*ex*_; NODDI’s *ficvf*; and SMI parameters *f*, 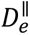 and 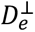 (Table 4; Fig. 4).

**Figure 4.**
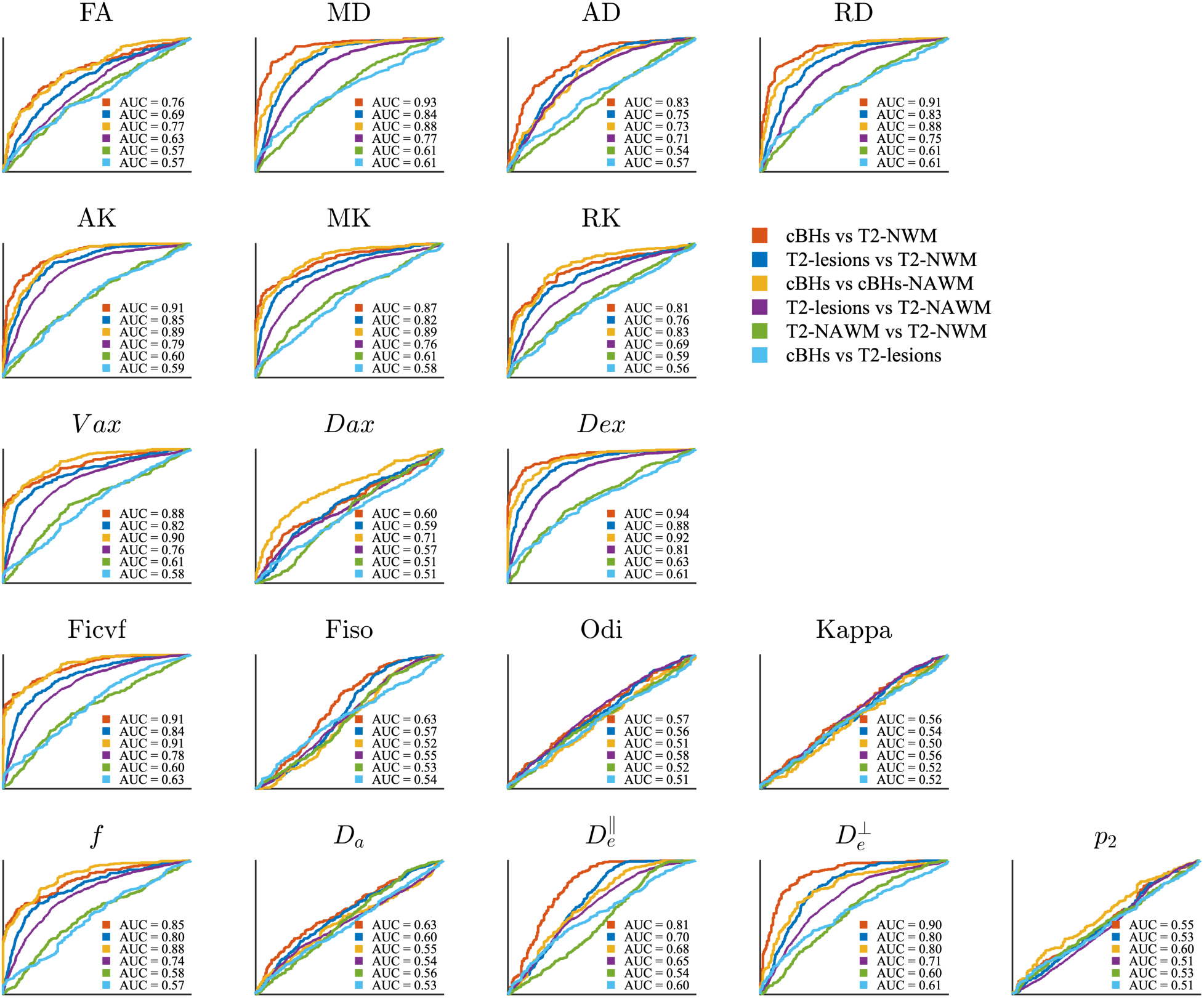
ROC analysis of all diffusion model parameters across matter tissue contrasts.

#### 3.3.2 Lesions and NAWM

Across lesion–NAWM contrasts, diffusion metrics from all models demonstrated meaningful discriminative performance and showed patterns largely consistent with those observed in the lesion vs T2-NWM comparisons. Specifically, high classification performance was observed for MD, AD, and RD in DTI; AK, MK, and RK in DKI; *V*_*ax*_ and *D*_*ex*_ in SMT; *ficvf i*n NODDI; and *f*, 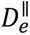 and 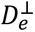 in SMI (Table 4; Fig. 4).

#### 3.3.3 NAWM and NWM

In the T2-NAWM versus T2-NWM contrast, diffusion metrics from all models demonstrated limited discriminative performance, with most AUC values hovering around 0.6. The highest AUC was observed for SMT’s *D*_*ex*_, with a value of 0.63 (Table 4; Fig. 4).

#### 3.3.4 cBHs and T2-lesions

In the cBHs versus T2-lesions contrast, diffusion metrics from all models demonstrated limited discriminative performance, with most AUC values around 0.6.

### 3.4 Discriminative Performance of Diffusion Models Across Tissue Classes

As shown in Figure 5, several tissue contrasts exhibited strong discriminative performance, with AUC values exceeding 0.8. In the cBHs vs T2-NWM comparison, the NODDI model achieved the highest discrimination (AUC = 0.98). Similarly, NODDI showed the best performance in distinguishing T2-lesions from T2-NWM (AUC = 0.90) and in cBHs vs cBHs-NAWM (AUC = 0.95). In contrast, discrimination between T2-NAWM and T2-NWM was generally modest across all models, and classification performance in the cBHs vs T2-lesions contrast was also limited, with the highest AUC observed for NODDI at 0.67.

**Figure 5.**
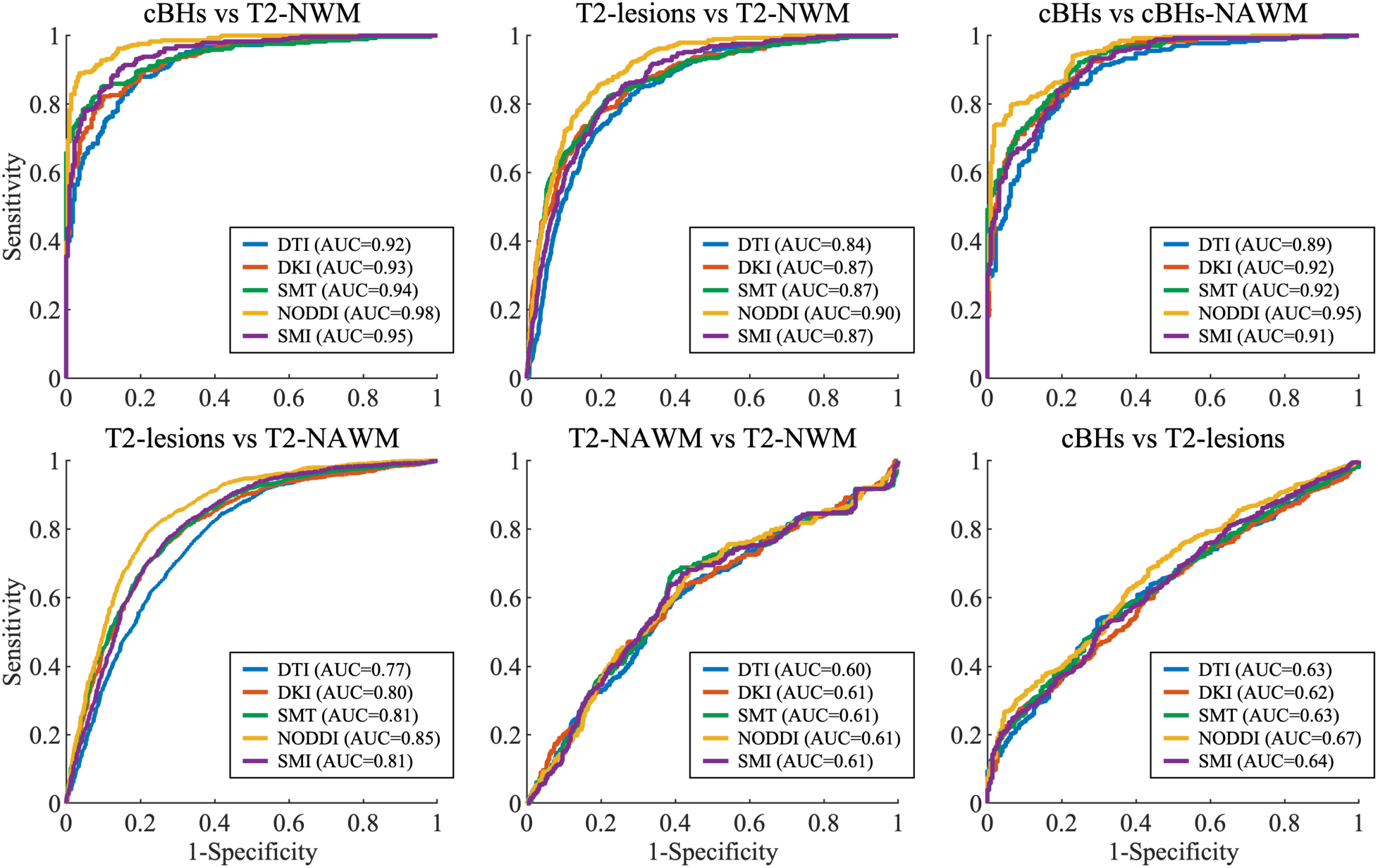
ROC curves for all evaluated diffusion models across white matter tissue contrasts. AUC values are shown in the legend.

### 3.5 Effect of Shell Strategy on the Discriminative Performance of Diffusion Models

As shown in Figure 6, classification performance generally improved from single-shell to two-shell models, with the joint model yielding the highest AUCs in most contrasts. For cBHs vs T2-NWM, AUC increased from 0.92 (single-shell) to 0.98 (two-shell) and 0.99 (joint). In T2-lesions vs T2-NWM, performance rose from 0.84 to 0.91 for both two-shell and joint models. Similar improvements were seen for cBHs vs cBHs-NAWM (0.89 → 0.95/0.96) and T2-lesions vs T2-NAWM (0.77 → 0.86/0.87). By contrast, in the subtler comparisons of T2-NAWM vs T2-NWM and cBHs vs T2-lesions, all strategies showed modest discrimination with AUCs around 0.60–0.67.

**Figure 6.**
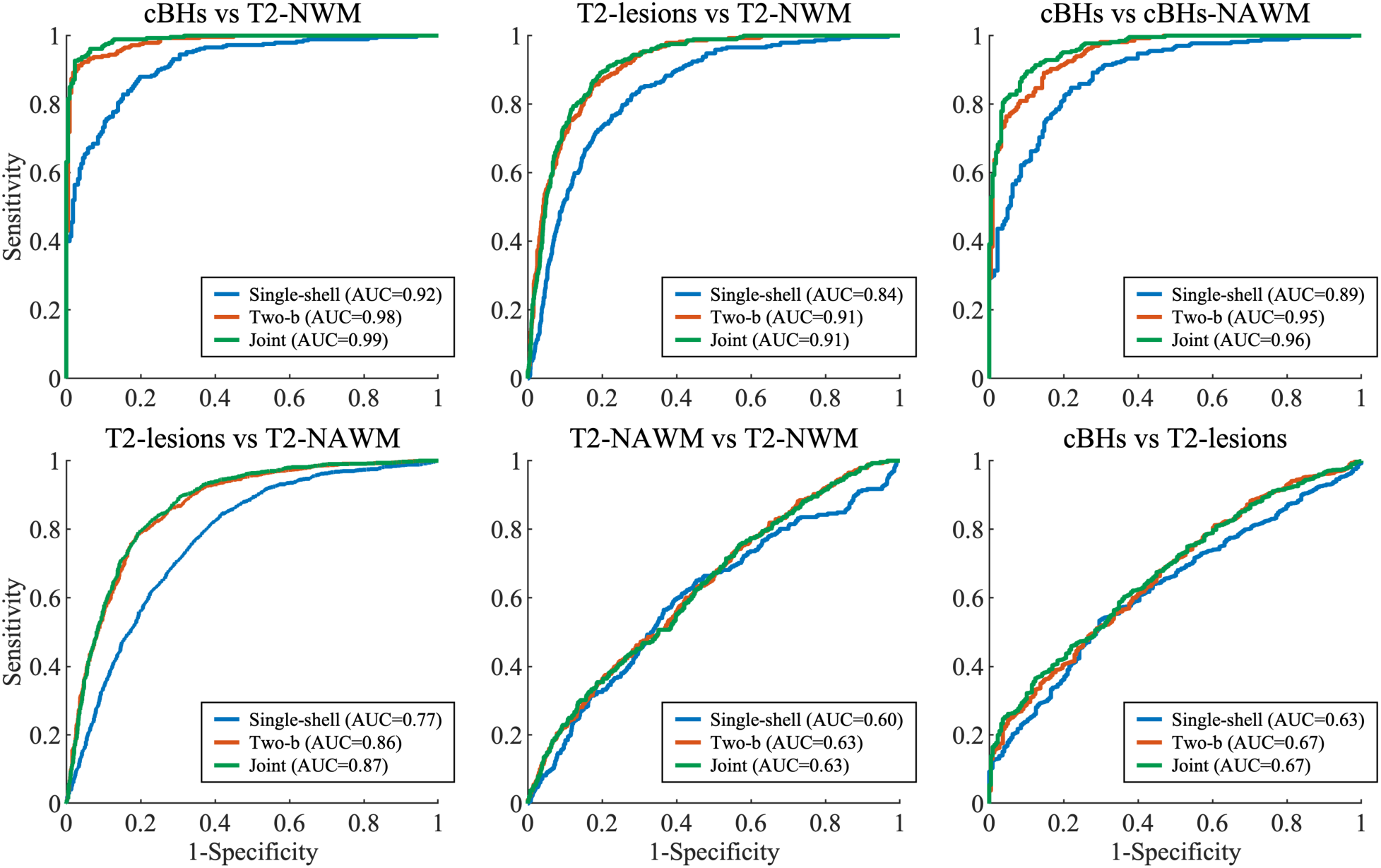
Effect of shell strategy on tissue contrast classification performance.

### 3.6 Identification of Key Discriminative Features in Joint Models Using LASSO

To identify the most informative diffusion features for tissue discrimination, LASSO regression was applied separately to each pairwise contrast, and features were ranked according to their importance. For cBHs vs T2-NWM, the top three ranked features were *ficvf*, RD, and *odi*; for T2-lesions vs T2-NWM, *odi*, *ficvf*, and DK ranked highest. In the cBHs vs cBHs-NAWM contrast, the top three features were *ficvf*, *odi*, and RD, whereas RD, *odi*, and *ficvf* were most informative for T2-lesions vs T2-NAWM. In the more subtle T2-NAWM vs T2-NWM comparison, *D*_*a*_, 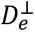 and *V_ax_* showed the highest rankings, while *ficvf*, 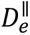, and 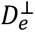 were identified as the top discriminative features for cBHs vs T2-lesions (Fig. 7).

**Figure 7.**
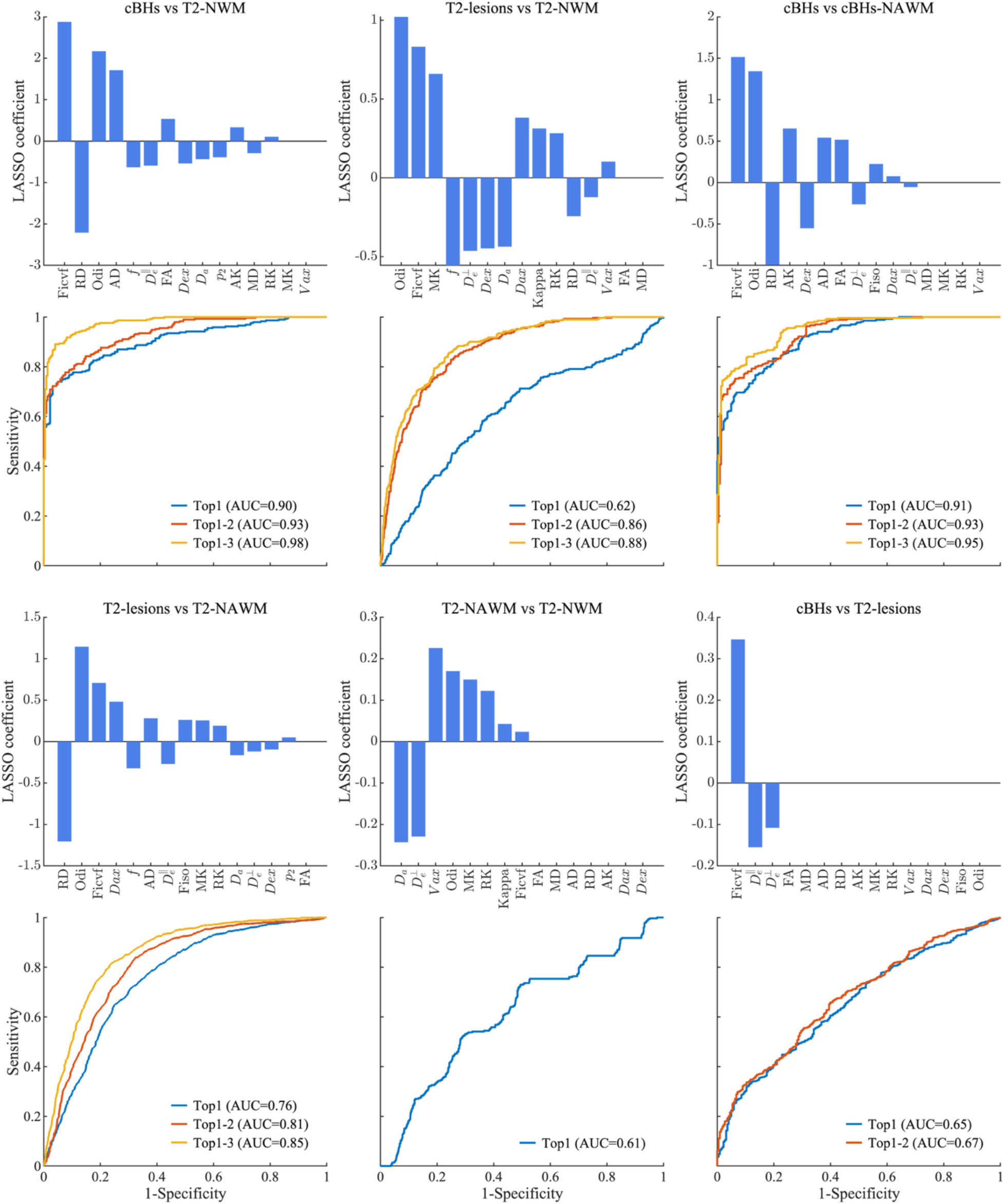
Top-ranked diffusion features and their incremental contribution to classification performance

These ranked features were subsequently evaluated using an incremental joint ROC analysis. ROC curves were generated by sequentially adding features in ranking order (Top-1, Top-2, Top-3, etc.), and the corresponding AUC values were assessed at each step. Feature inclusion was terminated when additional parameters no longer yielded meaningful improvement in classification performance. This strategy enabled identification of the smallest subset of features achieving stable or near-optimal AUC for each tissue contrast, as illustrated in Fig. 7.

## Discussion

In this study, we conducted a lesion-resolved, multi-model evaluation of diffusion MRI metrics across distinct white matter tissue classes in early MS using a large set of manually delineated ROIs. Across all diffusion models, lesions exhibited coherent microstructural alterations relative to normal white matter, characterized by reduced anisotropy, increased diffusivity, and decreased measures related to tissue complexity and axonal integrity, indicating that MS-related damage manifests along shared microstructural dimensions detectable by diffusion MRI. While many individual parameters showed strong discriminative performance for lesion–normal tissue contrasts, no single metric fully captured the spectrum of tissue alterations across classes; instead, higher-order diffusion models provided complementary discriminative information beyond conventional DTI. Importantly, these gains were strongly dependent on diffusion acquisition strategy, with classification performance improving systematically from single shell to multi-shell and joint modeling approaches. In contrast, more subtle tissue differences, such as those between NAWM and NWM or between lesion subtypes, remained difficult to resolve, underscoring both the sensitivity limits of diffusion MRI and the biological continuity of these tissue classes. Together, these findings indicate that multi-b-shell acquisitions are necessary to fully exploit the discriminative potential of higher-order diffusion models for lesion-resolved and tissue-specific characterization in MS.

Across diffusion models, lesions exhibited the most pronounced microstructural abnormalities relative to normal white matter, providing a clear reference for interpreting more subtle tissue alterations. Comparisons involving NAWM revealed largely concordant patterns of diffusion changes relative to lesions, supporting the notion that MS-related tissue damage extends beyond focal plaques along a continuous pathological spectrum[37, 38]. This interpretation is further supported by prior studies reporting widespread diffusion abnormalities in NAWM, including increased MD, AD, and RD and reduced FA across multiple white matter regions[39, 40]. In addition, a spatial gradient of diffusion changes has been described, with more pronounced abnormalities in NAWM proximal to lesions and progressively milder alterations with increasing distance from focal plaques[41]. Histological evidence of demyelination and axonal transection extending beyond lesion boundaries further suggests that diffusion abnormalities in NAWM may reflect Wallerian degeneration and related downstream processes[42, 43]. Together, these observations provide a biological framework for our findings, in which NAWM differs significantly from normal white matter yet exhibits more subtle diffusion alterations than focal lesions[44].

Across both DTI- and multi-b-shell–based diffusion models, parameters consistently demonstrated strong discriminative ability between lesions and normal tissue, including both NAWM and NWM, indicating robust sensitivity to major MS-related microstructural disruption[45]. In contrast, classification performance was markedly reduced for comparisons involving more closely related tissue classes, such as cBHs versus T2-lesions and T2-NAWM versus T2-NWM, despite statistically significant differences. This pattern suggests that while diffusion MRI is highly sensitive to gross pathological alterations, microstructural features distinguishing lesion subtypes or subtly affected tissue compartments exhibit substantial biological overlap, limiting separability at the voxel or ROI level[8]. Similar observations have been reported in prior diffusion MRI studies of MS, in which lesion–normal tissue contrasts were readily discriminable, whereas distinctions among lesion phenotypes or between NAWM and normal white matter were more subtle and heterogeneous[46, 47].

Beyond model selection, our findings highlight the critical role of diffusion acquisition strategy in determining discriminative performance. Across multiple tissue contrasts, diffusion models fitted using two b-shell data consistently outperformed single-shell–based approaches, and the joint models combining single- and two-shell information achieved the highest AUCs. This improvement suggests that multi-shell acquisitions capture complementary microstructural information that cannot be recovered from single b-shell data alone, likely reflecting enhanced sensitivity to compartment-specific diffusion processes[16, 48]. Importantly, feature selection using LASSO further demonstrated that a limited subset of diffusion parameters accounted for most of the discriminative power across tissue contrasts[49]. These features predominantly reflected biologically meaningful properties related to axonal density, orientation organization, and extra-axonal diffusivity[50]. Together, these results indicate that multi-b-shell acquisition strategies are not merely advantageous but necessary for robust tissue characterization in MS, and that focusing on a small number of key diffusion features may facilitate more interpretable and clinically actionable assessments of tissue pathology.

## Limitation

Several limitations of this study should be acknowledged. First, although many manually delineated ROIs were analyzed, the number of subjects was relatively modest, which may limit subject-level inference and the generalizability of the findings. Second, all analyses were performed at the ROI level using cross-sectional data, precluding assessment of longitudinal microstructural evolution and limiting causal interpretation of diffusion-derived changes across tissue classes. Third, while statistically significant differences were detected across lesions, NAWM, and normal white matter for both DTI and higher-order diffusion models, discriminative performance remained limited for more subtle contrasts, such as NAWM versus NWM and cBHs versus T2-lesions. This likely reflects substantial biological overlap and heterogeneous pathology across these tissue compartments, which constrains separability even when multi-model and multi-shell information is available. Fourth, although multi-b-shell acquisitions and joint modeling strategies consistently improved classification performance relative to single-shell approaches, the observed gains were moderate rather than transformative. This suggests that while multi-shell diffusion MRI provides complementary microstructural information, additional advances—such as richer sampling of diffusion encoding, incorporation of alternative microstructural models, or integration with complementary imaging modalities—may be required to further enhance tissue-specific characterization. Future studies employing larger cohorts, longitudinal designs, and expanded diffusion acquisition strategies will be important for refining diffusion-based biomarkers of MS pathology and improving their translational relevance.

## Conclusion

In summary, this study provides a lesion-resolved, multi-model evaluation of diffusion MRI across distinct white matter tissue classes in early multiple sclerosis. By leveraging a large set of manually delineated ROIs, we demonstrate that diffusion MRI robustly captures a continuum of microstructural alterations spanning focal lesions, NAWM, and normal white matter. While conventional DTI remains highly sensitive to major lesion-related pathology, higher-order diffusion models offer complementary insights into compartment-specific microstructural changes that are not fully represented by tensor-based measures alone. Importantly, our results show that the discriminative utility of diffusion models is strongly influenced by acquisition strategy, with two b-shell and joint modeling approaches consistently outperforming single-shell analyses. Feature selection further indicates that a limited set of biologically meaningful diffusion metrics accounts for much of the observed discriminative power, highlighting opportunities for more parsimonious and interpretable biomarker development. Together, these findings underscore the necessity of multi-b-shell diffusion acquisitions and multi-model analytical frameworks for robust, tissue-specific characterization of MS pathology, and provide practical guidance for optimizing diffusion MRI protocols in both research and clinical settings.

## Data Availability

All data produced in the present study are available upon reasonable request to the authors

## Acknowledgments

We are grateful to our patients and their families, and all the healthy controls who agreed to participate in this study. We thank Mr. Reece Clarke and Mr. Keejin Yoon for their invaluable help in setting up the study and start the recruitment process along with all the MRI technicians of the Vanderbilt University Institute of Imaging Science for assistance with scanning. We would like to express our gratitude to Dr. Harold Moses for his support with recruitment. This manuscript is dedicated to the sweet memory of Mr. Oscar Castr

## References

1. Jakimovski, D., et al., Multiple sclerosis. The Lancet, 2024. 403(10422): p. 183–202.

2. Portaccio, E., et al., Multiple sclerosis: emerging epidemiological trends and redefining the clinical course. The Lancet Regional Health – Europe, 2024. 44.

3. Kuhlmann, T., et al., Acute axonal damage in multiple sclerosis is most extensive in early disease stages and decreases over time. Brain, 2002. 125(10): p. 2202–2212.

4. Neema, M., et al., MRI in multiple sclerosis: what’s inside the toolbox? Neurotherapeutics, 2007. 4(4): p. 602–17.

5. Filippi, M., et al., Magnetic resonance techniques in multiple sclerosis: the present and the future. Arch Neurol, 2011. 68(12): p. 1514–20.

6. Sicotte, N.L., Neuroimaging in multiple sclerosis: neurotherapeutic implications. Neurotherapeutics, 2011. 8(1): p. 54–62.

7. Riva, M., et al., Tissue-specific imaging is a robust methodology to differentiate in vivo T1 black holes with advanced multiple sclerosis-induced damage. AJNR Am J Neuroradiol, 2009. 30(7): p. 1394–401.

8. Muñoz González, G., et al., A focus on the normal-appearing white and gray matter within the multiple sclerosis brain: a link to smoldering progression. Acta Neuropathol, 2025. 150(1): p. 16.

9. Elahi, R., et al., Advanced MRI Methods for Diagnosis and Monitoring of Multiple Sclerosis (MS). J Magn Reson Imaging, 2025. 62(6): p. 1546–1578.

10. Le Bihan, D., et al., Diffusion tensor imaging: concepts and applications. J Magn Reson Imaging, 2001. 13(4): p. 534–46.

11. Cassol, E., et al., Diffusion tensor imaging in multiple sclerosis: a tool for monitoring changes in normal-appearing white matter. Mult Scler, 2004. 10(2): p. 188–96.

12. Ge, Y., M. Law, and R.I. Grossman, Applications of diffusion tensor MR imaging in multiple sclerosis. Ann N Y Acad Sci, 2005. 1064: p. 202–19.

13. Caranova, M., et al., A systematic review of microstructural abnormalities in multiple sclerosis detected with NODDI and DTI models of diffusion-weighted magnetic resonance imaging. Magnetic Resonance Imaging, 2023. 104: p. 61–71.

14. Huang, S., et al., White Matter Abnormalities and Cognitive Deficit After Mild Traumatic Brain Injury: Comparing DTI, DKI, and NODDI. Front Neurol, 2022. 13: p. 803066.

15. Jelescu, I.O. and E. Fieremans, *Chapter 2 - Sensitivity and specificity of diffusion MRI to neuroinflammatory processes*, in *Advances in Magnetic Resonance Technology and Applications*, C. Laule and J.D. Port, Editors. 2023, Academic Press. p. 31–50.

16. Li, C.X., S. Patel, and X. Zhang, Evaluation of multi-shell diffusion MRI acquisition strategy on quantitative analysis using multi-compartment models. Quant Imaging Med Surg, 2020. 10(4): p. 824–834.

17. Ning, L., et al., Cross-scanner and cross-protocol multi-shell diffusion MRI data harmonization: Algorithms and results. NeuroImage, 2020. 221: p. 117128.

18. Jha, R.R., et al., Single-shell to multi-shell dMRI transformation using spatial and volumetric multilevel hierarchical reconstruction framework. Magnetic Resonance Imaging, 2022. 87: p. 133– 156.

19. Pines, A.R., et al., Leveraging multi-shell diffusion for studies of brain development in youth and young adulthood. Developmental Cognitive Neuroscience, 2020. 43: p. 100788.

20. Wu, E.X. and M.M. Cheung, MR diffusion kurtosis imaging for neural tissue characterization. NMR Biomed, 2010. 23(7): p. 836–48.

21. Zhang, H., et al., NODDI: practical in vivo neurite orientation dispersion and density imaging of the human brain. Neuroimage, 2012. 61(4): p. 1000–1016.

22. Henriques, R.N., S.N. Jespersen, and N. Shemesh, Microscopic anisotropy misestimation in spherical-mean single diffusion encoding MRI. Magnetic resonance in medicine, 2019. 81(5): p. 3245–3261.

23. Coelho, S., et al., Reproducibility of the standard model of diffusion in white matter on clinical MRI systems. NeuroImage, 2022. 257: p. 119290.

24. Shi, Z., et al., Microstructural alterations in different types of lesions and their perilesional white matter in relapsing-remitting multiple sclerosis based on diffusion kurtosis imaging. Multiple Sclerosis and Related Disorders, 2023. 71.

25. Seyedmirzaei, H., et al., Neurite Orientation Dispersion and Density Imaging in Multiple Sclerosis: A Systematic Review. J Magn Reson Imaging, 2023. 58(4): p. 1011–1029.

26. Bagnato, F., et al., Multi-compartment Spherical Microscopic Diffusion Imaging Using Spherical Mean Techniques to Probe Axonal Injury in Multiple Sclerosis (P3.383). Neurology, 2018. 90(15_supplement): p. P3.383.

27. Liao, Y., et al., Mapping tissue microstructure of brain white matter in vivo in health and disease using diffusion MRI. Imaging Neuroscience, 2024. 2.

28. Kurtzke, J.F., Rating neurologic impairment in multiple sclerosis: an expanded disability status scale (EDSS). Neurology, 1983. 33(11): p. 1444–52.

29. Feys, P., et al., The Nine-Hole Peg Test as a manual dexterity performance measure for multiple sclerosis. Mult Scler, 2017. 23(5): p. 711–720.

30. Kaufman, M., D. Moyer, and J. Norton, The significant change for the Timed 25-foot Walk in the multiple sclerosis functional composite. Multiple Sclerosis Journal, 2000. 6(4): p. 286–290.

31. Benedict, R.H., et al., Validity of the minimal assessment of cognitive function in multiple sclerosis (MACFIMS). Journal of the International Neuropsychological Society, 2006. 12(4): p. 549–558.

32. Stegen, S., et al., Validity of the California Verbal Learning Test–II in multiple sclerosis. The Clinical Neuropsychologist, 2010. 24(2): p. 189–202.

33. Tournier, J.-D., et al., MRtrix3: A fast, flexible and open software framework for medical image processing and visualisation. Neuroimage, 2019. 202: p. 116137.

34. Jenkinson, M., et al., Fsl. Neuroimage, 2012. 62(2): p. 782–790.

35. Bagnato, F., et al., Selective inversion recovery quantitative magnetization transfer imaging: Toward a 3 T clinical application in multiple sclerosis. Multiple Sclerosis Journal, 2020. 26(4): p. 457–467.

36. Oberg, A.L. and D.W. Mahoney, Linear mixed effects models. Topics in biostatistics, 2007: p. 213–234.

37. Filippi, M., et al., Multiple sclerosis. Nature Reviews Disease Primers, 2018. 4(1): p. 43.

38. Cree, B.A.C., et al., Secondary Progressive Multiple Sclerosis. Neurology, 2021. 97(8): p. 378–388.

39. Sbardella, E., et al., DTI Measurements in Multiple Sclerosis: Evaluation of Brain Damage and Clinical Implications. Mult Scler Int, 2013. 2013: p. 671730.

40. Preziosa, P., et al., Intrinsic damage to the major white matter tracts in patients with different clinical phenotypes of multiple sclerosis: a voxelwise diffusion-tensor MR study. Radiology, 2011. 260(2): p. 541–50.

41. Guo, A.C., V.L. Jewells, and J.M. Provenzale, Analysis of normal-appearing white matter in multiple sclerosis: comparison of diffusion tensor MR imaging and magnetization transfer imaging. AJNR Am J Neuroradiol, 2001. 22(10): p. 1893–900.

42. Kolasinski, J., et al., A combined post-mortem magnetic resonance imaging and quantitative histological study of multiple sclerosis pathology. Brain, 2012. 135(Pt 10): p. 2938–51.

43. Evangelou, N., et al., Regional axonal loss in the corpus callosum correlates with cerebral white matter lesion volume and distribution in multiple sclerosis. Brain, 2000. 123 ( **Pt 9**): p. 1845–9.

44. Toubasi, A.A., et al., Improving the Assessment of Axonal Injury in Early Multiple Sclerosis. Academic Radiology, 2025. 32(2): p. 1002–1014.

45. Hagiwara, A., et al., White Matter Abnormalities in Multiple Sclerosis Evaluated by Quantitative Synthetic MRI, Diffusion Tensor Imaging, and Neurite Orientation Dispersion and Density Imaging. AJNR Am J Neuroradiol, 2019. 40(10): p. 1642–1648.

46. Moll, N.M., et al., Multiple sclerosis normal-appearing white matter: pathology-imaging correlations. Ann Neurol, 2011. 70(5): p. 764–73.

47. Vrenken, H., et al., Normal-appearing white matter changes vary with distance to lesions in multiple sclerosis. AJNR Am J Neuroradiol, 2006. 27(9): p. 2005–11.

48. Vanden Bulcke, C., et al., Comparative overview of multi-shell diffusion MRI models to characterize the microstructure of multiple sclerosis lesions and periplaques. NeuroImage: Clinical, 2024. 42: p. 103593.

49. Bose, G., et al., Early Predictors of Clinical and MRI Outcomes Using Least Absolute Shrinkage and Selection Operator (LASSO) in Multiple Sclerosis. Ann Neurol, 2022. 92(1): p. 87–96.

50. Cerdán Cerdá, A., et al., A translational MRI approach to validate acute axonal damage detection as an early event in multiple sclerosis. Elife, 2024. 13.

